# Clinical prediction rule to guide diagnostic testing for *Shigellosis* and improve antibiotic stewardship for pediatric diarrhea

**DOI:** 10.1101/2022.12.19.22283701

**Authors:** Sharia M. Ahmed, Ben J. Brintz, Patricia B. Pavlinac, Md Iqbal Hossain, Ashraful Islam Khan, James A. Platts-Mills, Karen L. Kotloff, Daniel T. Leung

## Abstract

**Background:** Diarrheal diseases are a leading cause of death for children under-5.

Identification of etiology helps guide pathogen-specific therapy, but availability of diagnostic testing is often limited in low resource settings. Our goal is to develop a clinical prediction rule (CPR) to guide clinicians in identifying when to use a point-of-care diagnostic for *Shigella* in children presenting with acute diarrhea.

**Methods:** We used clinical and demographic data from the Global Enteric Multicenter Study (GEMS) study to build predictive models for diarrhea of *Shigella* etiology in children ≤59 months presenting with moderate-to-severe diarrhea in Africa and Asia. We screened variables using random forests, and assessed predictive performance with random forest regression and logistic regression using cross-validation. We used the Etiology, Risk Factors, and Interactions of Enteric Infections and Malnutrition and the Consequences for Child Health and Development (MAL-ED) study to externally validate our GEMS-derived CPR.

**Results:** Of the 5011 cases analyzed, 1332 (27%) had diarrhea of *Shigella* etiology. Our CPR had high predictive ability (AUC=0.80 (95% CI: 0.79, 0.81) using the top two predictive variables, age and caregiver reported bloody diarrhea. We show that by using our CPR to triage who receives diagnostic testing, 3 times more *Shigella* diarrhea cases would have been identified compared to current symptom-based guidelines, with only 27% of cases receiving a point-of-care diagnostic test.

**Conclusions:** We demonstrate how a clinical prediction rule can be used to guide use of a point-of-care diagnostic test for diarrhea management. Using our CPR, available diagnostic capacity can be optimized to improve appropriate antibiotic use.

**Key points:** Using an externally validated clinical prediction tool to triage who receives diagnostic testing, 3 times more *Shigella* diarrhea cases would have been identified compared to current symptom-based guidelines, with only 27% of cases receiving a point-of-care diagnostic test.

## INTRO

Despite medical advancements, diarrheal diseases remain a leading cause of death in children under-5, with an estimated 960 million cases and 500,000 deaths worldwide, most of which occur in low and middle-income countries (LMICs). While most episodes of diarrhea are self-limiting and treatment with oral rehydration adequate, antibiotics can reduce the severity and duration of some diarrhea etiologies, including *Shigella* spp, the cause of shigellosis[1, 2]. Therefore, the World Health Organization’s (WHO) 2014 updated Integrated Management of Childhood Illness (IMCI) recommends antibiotics for treatment of presumptive *Shigella spp* infection with severe diarrhea[3, 4]. In practice, antibiotics may also be administered for presumptive shigellosis if the patient is malnourished, even in the absence of IMCI-indicated diarrhea-associated symptoms. Conversely, antibiotics can be expensive, and cause adverse effects, including hypersensitivity reactions, nausea, and prolonged symptoms for other diarrhea etiologies[5], and overuse of antibiotics can contribute to antimicrobial resistance[6–12]. Given changing resistance patterns, diarrhea etiology can inform antibiotic choice. Therefore, accurately identifying the etiology or cause of diarrhea is vital for proper management at the individual level and appropriate public health responses at the population level.

Laboratory-based diagnostics such as stool culture and PCR remain the most commonly used methods of identifying diarrhea etiology, including *Shigella spp[13–15]*. Unfortunately, currently available laboratory diagnostics are time and resource intensive, and thus are rarely utilized for diarrheal case management in LMICs. Therefore, the IMCI recommends using the nature of diarrhea stool to determine etiology, with blood in stool (“dysentery”) indicative of *Shigella* infection[3]. However, the accuracy of dysentery as an indicator of *Shigella* infection can vary widely[16], with up to 90% of shigellosis cases remaining undiagnosed in a recent study[17].

Advances in diagnostic methods, including immunochromatographic dipsticks and the loop-mediated isothermal amplification (LAMP) platform, offer hope for expanded diagnostic testing capacity in low-resource settings[18, 19]. These technologies aim to provide rapid, point-of-care (POC) diagnostic testing capacity that can provide etiologic information in a fraction of the time of existing methods, without the need for specialized laboratory resources, personnel, or cold chains[20]. However, while such diagnostic tests would likely be cheaper than existing technologies, it is still unlikely that POC tests will be available to every patient presenting to care with diarrhea in LMICs.

In the absence of universally available, affordable, comprehensive testing for diarrhea etiology, clinical prediction rules (CPRs) offer a mechanism to optimize the usage of available diagnostic testing capability and financing. CPRs are algorithms that guide clinicians in clinical decision making, and are well-accepted in other areas of medicine[21]. The goal of this study was to develop clinical prediction tools to identify patients for whom definitive diagnosis would change treatment, in order to guide clinicians in identifying when to use a hypothetical POC test for making diarrhea treatment decisions. We then estimated the accuracy of this proposed testing regimen compared to current guidelines.

## METHODS

### Study Population for Derivation Cohort (GEMS)

We derived CPRs to predict *Shigella* infection using data from The Global Enteric Multicenter Study (GEMS). GEMS has been described previously[13, 22]. In summary, GEMS was a prospective case-control study of acute moderate to severe diarrhea (MSD) in children 0-59 months of age from 7 sites in Africa and Asia from December 2007 – March 2011. At initial presentation to a sentinel hospital or health center, diarrhea cases were enrolled and matched within 14 days to 1-3 diarrhea-free community controls. Only data from diarrhea cases were used in this study. Diarrhea was defined as new onset (after ≥7 days diarrhea-free) of 3 or more looser than normal stools in the previous 24 hours lasting 7 days or less, and MSD was defined as diarrhea plus one or more of the following: dysentery (blood in stool), dehydration, or hospital admission. Caregivers provided demographics, epidemiological, and clinical information via standardized questionnaires. Clinic staff conducted physical exams and collected stool samples which have undergone molecular testing.

Participants’ parents or caregivers provided informed consent, either in writing or witnessed if parents/caregivers were illiterate. The GEMS study protocol was approved by ethical review boards at each field site and the University of Maryland, Baltimore, USA.

### Study Population for Validation Cohort (MAL-ED)

We externally validated our CPR using data from the Etiology, Risk Factors, and Interactions of Enteric Infections and Malnutrition and the Consequences for Child Health and Development (MAL-ED) study. Extensive study details of MAL-ED have been described elsewhere[23–26]. In brief, MAL-ED is a longitudinal birth cohort of from October 2009 – March 2012 in 8 countries in Africa and Southeast Asia. Healthy children were enrolled within 17 days of birth and prospectively followed through 24 months of age. Information on household, demographic, and clinical data from mother and child were collected at enrollment and reassessed periodically. Households were visited twice-weekly, and stool samples were collected from children with diarrhea, defined as maternal report of three or more loose stools in a 24 hour period, or one loose stool with blood. Each distinct diarrhea episode was separated by at least 2 days without symptoms. Stools then underwent molecular testing to ascertain etiology[25].

Informed consent was obtained from parents/caregivers of participants. The MAL-ED study protocol was approved by ethical review boards at each field site, the University of Virginia Institutional Review Board for Health Sciences Research, Charlottesville, USA, and the Johns Hopkins Institutional Review Board, Baltimore, USA.

### Outcomes

Our outcome of interest was diarrhea caused by *Shigella* infection. We used the quantitative real-time qPCR attribution models developed by Liu et. al. to assign diarrhea episode-specific attributable fractions (AFe) of etiology for each diarrhea episode[27]. We considered AFe of *Shigella* ≥ 0.5 as *Shigella* attributable diarrhea [24].

### Predictive Variables

We explored over 130 potential predictors collected at enrollment in GEMS, including descriptors of the child, household, and community (Supplementary Table S1). We did not consider composite variables (e.g. wealth index) since their utilization in the final CPRs would require collecting multiple variables already considered individually.

### Development of the Clinical Prediction Rule

First, we screened possible predictors using random forests. An ensemble learning method, random forests builds multiple decision trees (1000 throughout this analysis) on bootstrapped samples of the data. Variability is reduced and the trees are decorrelated because only a random sample of potential predictors are considered at each split[28]. We ranked the predictive importance of variables via the reduction in mean squared prediction error achieved by including the variable in the predictive model on out-of-bag samples (i.e. observations not in the bootstrapped sample).

Second, we used repeated cross-validation to assess internal model discrimination and generalizable performance. Random forests were fit to a training dataset consisting of a random 80% sample of the analytic dataset, and variables importance was ranked as above. This process was repeated over 100 iterations. We fit separate logistic and random forest regression models to the top predictive variables in the training dataset, examining the top 1-10, 15, 20, 30, 40, and 50 predictors. We then predicted the outcome (*Shigella* attributable diarrhea) on the test dataset for each iteration. We used the receiver operating characteristic (ROC) curve and the C-statistic (area under the ROC curve (AUC)) from the cross-validation to assess model discrimination.

Third, we assessed model calibration. Calibration refers to a model’s ability to correctly estimate the risk of the outcome[29, 30]. We assessed calibration intercept, or calibration-in-the-large, by modeling the log-odds of the true status, offset by the CPR-predicted log-odds. Next, we fit a logistic regression model with the CPR-predicted log-odds as the independent variable and the log-odds of the true status as the dependent variable to assess calibration slope. Finally, we graphically assessed moderate calibration by plotting the mean predicted probability of *Shigella* attributable diarrhea by the observed proportions (see Supplement for details and https://github.com/LeungLab/ShigellaDxStewardshipCPR for full code).

### Sensitivity and Subgroup Analyses

We performed multiple sensitivity analyses. First, we explored the relative discriminative performance of markers of malnutrition and child growth (MUAC, HAZ, or both). Second, we explored the age-strata specific CPRs, for children 0-11months, 12-23months, and 24-59 months. Third, we examined alternative definitions of *Shigella* etiology, namely AFe ≥ 0.3 and AFe ≥ 0.7. Fourth, we fit the CPR to only cases of bloody diarrhea, and only cases of non-bloody diarrhea. Fifth, we fit the CPR to all observations from all study sites except Bangladesh (due to its outlier proportion of *Shigella* etiology, see Table S2). Sixth, we added variables for stool descriptors as observed by clinicians (original stool descriptors were caregiver report at any time during the diarrhea episode). Seventh, we included an indicator variable for season (April-September versus October-March). Finally, we fit country-specific models, and conducted a quasi-external validation within the GEMS data by fitting a model to one continent and validating it on the other.

### External Validation: Estimating the Potential Impact of a CPR-Guided Diagnostic Testing Regimen in a New Population

In order to assess the potential impact of our final CPR on clinical practice, we evaluated how use of our CPR in a *CPR-guided diagnostic testing regimen* would have changed accuracy of presumptive diagnosis and appropriate antibiotic care in the presence of non-universal POC test availability. We used the CPR derived in GEMS data, and applied it to the MAL-ED population (external validation). We assumed every child presenting to care for diarrhea was screened using the CPR, resulting in a predicted probability that child’s diarrhea was of *Shigella* etiology. Those children with predicted probability of *Shigella* attributable diarrhea below cutoff x were deemed “not presumptive *Shigella*” and did not receive additional diagnostic testing or antibiotic treatment. Children with predicted probability of *Shigella* attributable diarrhea above cutoff y were deemed “presumptive *Shigella*” and received antibiotic treatment appropriate to their presumed infection, without additional diagnostic testing. Children whose CPR-predicted probability of *Shigella* attributable diarrhea was indeterminate (between cutoff x and y) received the POC diagnostic test, and results of the POC test determined diagnosis and treatment (see Supplemental Figure S1). We varied the range of cutoffs x and y, and compared the accuracy (e.g. sensitivity, specificity, etc.) of this CPR-guided diagnostic testing regimen to the current guidelines of presence/absence of dysentery for presumptive *Shigella* diagnosis. Finally, we varied the sensitivity and specificity of the POC test between 0.8 and 1.0 to explore the POC test’s potential impact on CPR-testing regimen accuracy.

## RESULTS

### Shigellosis in children presenting to care for acute diarrhea in GEMS

9439 children with acute diarrhea were enrolled in GEMS. Of these, 5304 had an attributable etiology. 17 observations were dropped for having unrealistic HAZ scores (<-7 or >7), and an additional 276 observations were dropped for having missing predictor data. This leaves an analytic sample size of 5011, of which 1332 (26.6%) had diarrhea due to *Shigella* etiology (AFe of *Shigella* ≥ 0.5) (Supplemental Figure S2). The proportion of diarrhea caused by *Shigella* etiology varied by location. *Shigella* was the most common etiology in Bangladesh, with 496 out of 876 (56.6%) diarrhea cases being attributable to *Shigella*. In contrast, Mozambique and Kenya had a very low proportion of diarrhea cases attributable to *Shigella*, only 20% and 13%, respectively (Supplemental Table S2). The proportion of diarrhea attributable to *Shigella* also varied by children’s age. Older children were most likely to have diarrhea attributable to *Shigella*, with 37%, 35%, and 9% of diarrhea being attributable to *Shigella* in children 24-59 months (mo), 12-23mo, and 0-11mo, respectively in GEMS. This pattern was similar in MAL-ED (Supplemental Table S3).

### Derivation of a CPR to identify children likely to have *Shigella* etiology amongst children presenting with acute diarrhea in GEMS data

During random forest screening of variables, logistic regression and random forest regression produced similar AUCs (Supplementary Figure S3), therefore we only present the easier to interpret logistic regression results. Table 1 presents the top-10 variables most predictive of *Shigella* etiology, ranked from most to least important. The top 10 variables were: age, caregiver reported blood in stool, MUAC, respiratory rate, temperature, sunken eyes, number of people living in the household, site, number of days of diarrhea at presentation, and number of people sleeping in the household. A model with 20 variables produced a maximum average cross-validated (cv)AUC of 0.81 (95% CI: 0.80, 0.82), while an cvAUC of 0.80 (95% CI: 0.79, 0.81), 0.79 (95% CI: 0.78, 0.8), and 0.79 (95% CI: 0.78, 0.80) was obtained with a CPR of 2, 5, and 10 variables, respectively (Supplementary Figure S3). We achieved a specificity of 0.42 at a sensitivity of 0.8 for the main 2-variable model (Figure 1). The average predicted probability of diarrhea of *Shigella* etiology was consistently close to the average observed probability (calibration intercept, or calibration-in-the-large), and the spread of predicted probabilities was similar to the spread of observed probabilities (calibration slope) for models including 1 to 10 predictor variables (Table 1, Figure 2, Supplemental Figure S5). Odds ratios for the 2-variable prediction model fit in GEMS are in Supplemental Table S4.

**Table 1:** Variable importance ordering, cross-validated average overall AUC, and AUC 95% confidence intervals for logistic regression model for predicting *Shigella* etiology (attributable fraction (AFe) of *Shigella* ≥ 0.5) in children <5 years old in 7 LMICs derived from GEMS data. Calibration intercepts and slopes.

| Number of predictor variables | GEMS-derived, performance in GEMS |  |  | GEMS-derived, performance in MAL-ED |  |
| --- | --- | --- | --- | --- | --- |
|  |  | Calibration Intercept (95% CI) | Calibration Slope (95% CI) | Calibration Intercept (95% CI) | Calibration Slope (95% CI) |
| AUCs – 2 var | 0.80 (0.8, 0.81) |  |  |  |  |
| 5 | 0.79 (0.79, 0.80) |  |  |  |  |
| 10 | 0.80 (0.80, 0.80) |  |  |  |  |
| 1 | Age (months) | -1.2 x10<sup>-2</sup><br>(-1.6 x10<sup>-1</sup>, 1.3 x10<sup>-1</sup>) | 1.00 (0.72, 1.29) |  |  |
| 2 | Caregiver reported blood in stool | -1.3 x10<sup>-2</sup><br>(-1.8 x10<sup>-1</sup>, 1.5 x10<sup>-1</sup>) | 1.00 (0.86, 1.14) | -0.02 (-0.20, 0.14) | 1.03 (0.80, 1.26) |
| 3 | MUAC | -1.3 x10<sup>-2</sup><br>(-1.8 x10<sup>-1</sup>, 1.5 x10<sup>-1</sup>) | 1.00 (0.86, 1.14) |  |  |
| 4 | Respiratory rate | -1.3 x10<sup>-2</sup><br>(-1.8 x10<sup>-1</sup>, 1.5 x10<sup>-1</sup>) | 1.00 (0.86, 1.14) |  |  |
| 5 | Temperature | -1.3 x10<sup>-2</sup><br>(-1.8 x10<sup>-1</sup>, 1.5 x10<sup>-1</sup>) | 0.99 (0.86, 1.13) |  |  |
| 6 | Sunken eyes | -1.2 x10<sup>-2</sup><br>(-1.8 x10<sup>-1</sup>, 1.5 x10<sup>-1</sup>) | 0.99 (0.86, 1.13) |  |  |
| 7 | Num. people living in household | -1.2 x10<sup>-2</sup><br>(-1.8 x10<sup>-1</sup>, 1.5 x10<sup>-1</sup>) | 0.99 (0.86, 1.13) |  |  |
| 8 | Site | -1.3 x10<sup>-2</sup><br>(-1.8 x10<sup>-1</sup>, 1.5 x10<sup>-1</sup>) | 0.99 (0.85, 1.13) |  |  |
| 9 | Num. days of diarrhea at presentation | $-1.4 \times 10^{-2}$<br>($-1.8 \times 10^{-1}$, $1.5 \times 10^{-1}$) | 0.99 (0.85, 1.12) | | |
| 10 | Num. rooms used for sleeping | $-1.3 \times 10^{-2}$<br>($-1.8 \times 10^{-1}$, $1.5 \times 10^{-1}$) | 0.98 (0.85, 1.16) | | |

**Figure 1:**
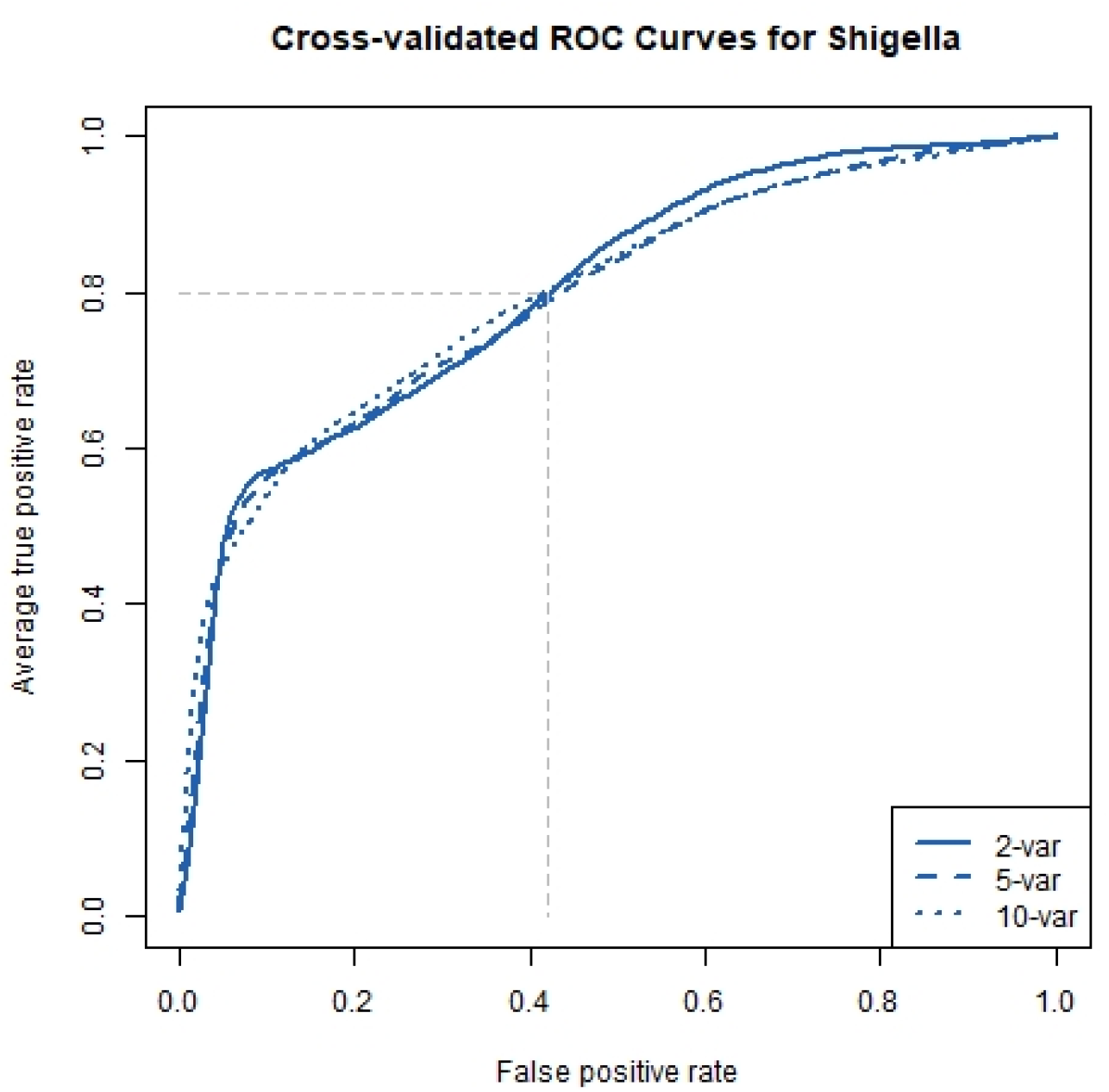
ROC curve: Average ROC curves from the internal cross-validated logistic regression models predicting *Shigella* etiology with 2, 5, and 10 predictors. The faded dashed lines represent specificity (1-false positive rate) achievable with a sensitivity (true positive rate) of 0.80 for prediction of the outcome

**Figure 2:**
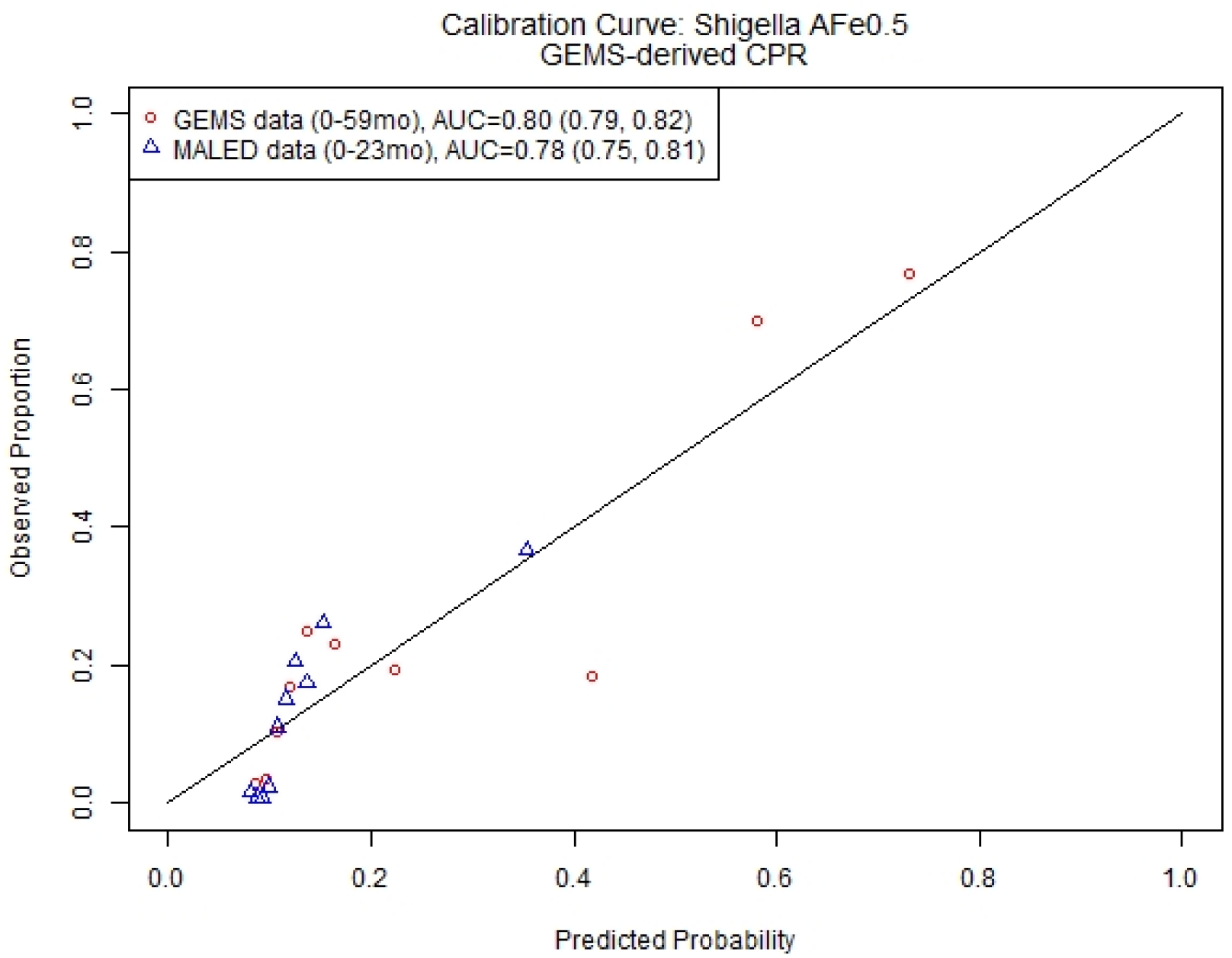
2-Variable CPR for *Shigella* etiology. Calibration curve and discriminative ability of 2-variable (age, presence of bloody diarrhea) model predicting *Shigella* etiology (attributable fraction (AFe) of *Shigella* ≥ 0.5) in children presenting for acute diarrhea in LMICs.

### Patient subsets and additional predictive variables did not meaningfully improve discriminative performance of CPR to identify diarrhea of *Shigella* etiology

The results of the sensitivity analyses are presented in Supplementary Table S5. Top predictors were highly consistent across all sensitivity analyses, with age, MUAC, and bloody diarrhea being top predictors in all relevant subgroups. While MUAC and HAZ were top predictors both individually and together, there was not a meaningful difference in the AUC compared to each other. Similarly, the CPR for the highest age strata 24-59months was not meaningfully different than the all-age model (0.80, 95% CI: 0.77, 0.82 for 24-59mo; 0.80, 95% CI: 0.79, 0.81 for 0-59mo), and the CPR for lower age strata had lower AUCs (0.75, 95% CI: 0.71, 0.78; 0.73, 95% CI: 0.71, 0.75 for 0-11mo and 12-23mo, respectively). Likewise, the CPRs using alternative attributable fraction cutoffs, additional clinician-observed stool description, and season all had AUCs around 0.80. The CPRs fit to subgroups of only bloody diarrhea cases, only non-bloody diarrhea cases, and excluding children in Bangladesh all resulted in lower AUCs (Supplementary Table S5).

### The derived CPR performed well at identifying diarrhea of *Shigella* etiology in an external population

Because there was only marginal improvements in discrimination (AUC) for additional variables beyond 2 predictors (Supplemental Figure S3), we elected to externally validate and explore the potential clinical impact of a 2-variable CPR. Therefore, we took the 2-variable CPR of *Shigella* etiology derived from children 0-59months of age in GEMS, including age and caregiver report of bloody diarrhea, and externally validated it in MAL-ED data. The CPR had good discrimination in GEMS (AUC=0.80, 95% CI: 0.79, 0.81), with a slight decrease in MAL-ED (AUC=0.77, 95% CI: 0.73, 0.81). On average, the CPR slightly overestimated the probability of diarrhea of *Shigella* etiology (calibration intercept −0.18, 95% CI: −0.36, −0.01), and prediction was slightly too extreme (calibration slope 0.93, 95% CI: 0.72, 1.14) (Table 1, Figure 2, Supplemental Figure S5). The model performed similarly when limiting the derivation dataset to children in GEMS age 0-23 months, with AUC=0.76 (95% CI: 0.75, 0.78) in GEMS, and AUC=0.77 (95% CI: 0.74, 0.81) at external validation in MAL-ED.

### A CPR-guided testing regimen for acute diarrhea could improve clinical care, even with decreased accuracy of point-of-care diagnostic tests

Using the 2-variable CPR derived in GEMS described above, we applied the CPR to MAL-ED data and explored how accurately our *CPR-guided diagnostic testing regimen* identified acute diarrhea patients with *Shigella* etiology, compared to the dysentery-based guidelines. Assuming a POC test sensitivity and specificity of 0.9 each, we found that the CPR-guided regimen would accurately identify more patients with *Shigella* etiology with as little as 10% of diarrhea patients receiving the POC test (sensitivity of regimen), compared to WHO IMCI-based dysentery-guided decision making (Figure 3). However, as the proportion of diarrhea patients receiving the POC test increased, there was an accompanying increase in patients being incorrectly identified as having diarrhea of *Shigella* etiology (false positive rate of regimen). By exploring the intersection of the sensitivity and false positive rate of the regimen, we identified an optimum CPR predicted probability range of 0.13 to 0.55. In other words, if all children in MAL-ED for whom the GEMS-derived CPR predicted probability of *Shigella* etiology was between 0.13 and 0.55 had received a POC diagnostic test, we would have conducted diagnostic testing on only 27% of pediatric diarrhea patients, and correctly identified 3 times more diarrhea cases of *Shigella* etiology, with no increase in the number of children incorrectly identified has having diarrhea of *Shigella* etiology, compared to the dysentery-guided determination.

**Figure 3:**
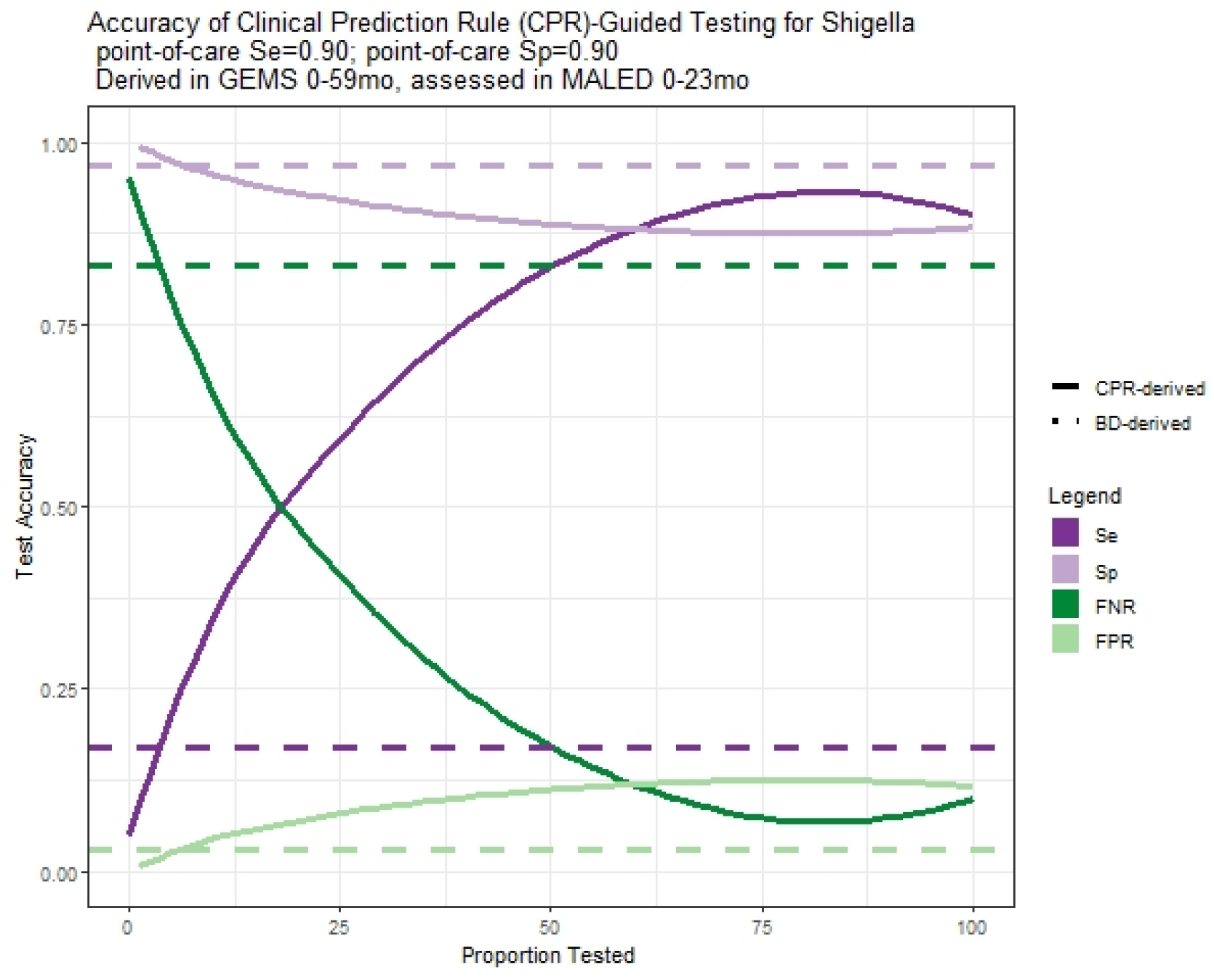
Performance of CPR-guided diagnostic testing regimen. CPR derived in GEMS, assessed in MAL-ED CPR=clinical prediction rule; BD=bloody diarrhea/dysentery; Se=sensitivity; Sp=specificity; FNR=false negative rate; FPR=false positive rate

Finally, we varied the POC test accuracy to estimate how this would have impacted the overall accuracy of the CPR-guided diagnostic testing regimen. We found that the sensitivity of the CPR-guided regimen was responsive to the sensitivity of the POC, with the regimen sensitivity decreasing slightly at lower POC test sensitivities. However, the CPR-guided regimen sensitivity was still much higher than the dysentery-guided sensitivity, even at lower POC test sensitivities. The regimen sensitivity did not vary with POC test specificity (see Supplemental Figure S6). The false positive rate of the CPR-guided regimen responded meaningfully to the specificity of the POC test, with regimen false positive rate increasing as POC test specificity decreased. Regimen false positive rate also increased slightly as POC test sensitivity decreased, but to a lesser extent. At most proportions of diarrhea patients tested, the CPR-guided regimen had a higher false positive rate than the dysentery-based guidelines, leading to more diarrhea patients incorrectly being diagnosed as having diarrhea of *Shigella* etiology.

## DISCUSSION

We derived and externally validated a clinical prediction rule for diarrhea of *Shigella* etiology using data from two large multi-site studies of pediatric diarrhea. Our CPR for *Shigella* etiology had good cross-validated discriminative performance in the derivation dataset (AUC=0.80, 95% CI: 0.79, 0.82, based on GEMS 0-59 months), and continued to perform well in the external dataset (AUC=0.77, 95% CI: 0.73, 0.81, based on MAL-ED 0-24 months). When hypothetically combined with emerging POC diagnostic tests, we demonstrate how a CPR-guided diagnostic testing regimen could lead to a 3-fold increase in the number of pediatric patients correctly diagnosed with, and therefore appropriately treated for, *Shigella* infection. By using our CPR to triage who receives a POC, we demonstrated how these gains in antibiotic stewardship could be achieved while still only testing a quarter of pediatric diarrhea patients.

There is increasing recognition of the limitations of dysentery as an indicator of *Shigella* infection (low sensitivity). A 2017 systematic review found that the sensitivity of dysentery to identify *Shigella* infection ranged from 1.9 – 85.9%[16]. This is consistent with what we observed in GEMS, where the sensitivity of caregiver reporter dysentery was only 17%.

Furthermore, they found that, on average, this sensitivity has decreased over time. Possible explanations include changes in circulating strains of *Shigella*, or in profiles of comorbidities and coinfections. They note that while *Shigella* infection is associated with death, dysentery is not associated with death, but also lack of dysentery is not indicative of a low mortality risk. Together, this highlights the need for improved diagnostic guidelines.

Current dysentery-based recommendations do provide guidance and are realistic given the prevalence of pediatric diarrhea and the limited availability of diagnostic testing resources in LMICs. Emerging technologies, including POC tests, have the potential to vastly improve our ability to decrease inappropriate use of antibiotics while simultaneously reducing *Shigella* morbidity and mortality by appropriately providing antibiotics to more *Shigella* patients.

However, the financial cost and consistent availability of POC tests in these settings have yet to be seen. We provide a way to achieve meaningful improvements in patient care with modest (and hopefully achievable) expansions in diagnostic testing.

Our CPR-guided diagnostic testing regimen also has the potential to lead to more equitable, standardized medical care. Another recent, large, multi-site study of young children with acute illness in similar locations found that 87% of diarrhea cases were prescribed antibiotics without a documented indication[31]. Given the poor sensitivity of current dysentery-based guidelines[17], it is unsurprising that clinicians rely on their clinical gestalt when making antibiotic treatment decisions. It is also possible that symptom extrinsic factors, such as caregivers’ desire for medication or household financial circumstances, are major contributors to treatment decisions[32]. Our CPR-guided diagnostic testing regimen provides a standardized, evidence-based strategy to guide antibiotic treatment decisions for pediatric diarrhea, with much greater accuracy than current practice.

Our study has a number of strengths and weaknesses. We derived a CPR for diarrhea of *Shigella* etiology from a multi-site, prospective study with extensive etiologic testing. While our derivation dataset included patients from a wider age range than our external validation dataset, (0-59months in GEMS, 0-23months in MAL-ED), our CPR had similarly high discriminative performance in subgroup analysis of more limited age groups. Furthermore, our quasi-external validation between continents within GEMS had similar top predictors and discriminative performance, as did our country-specific models. Similarly, we explored a range of AFe cutoffs for *Shigella* etiology, with consistent results. While our complete-case analysis strategy could introduce bias due to missing data, our CPR performed very similarly on an external dataset unlikely to have the same patterns of missingness. Finally, we demonstrated how our CPR could guide diagnostic stewardship. We assumed a POC test sensitivity and specificity of 0.9 each.

These are conservative estimate, as emerging technologies have reported higher test accuracies[20]. Our sensitivity analyses of POC test accuracy indicate that improved POC test accuracy would only improve our overall *Shigella* diagnostic accuracy.

In conclusion, we derived and externally validated a clinical prediction rule for diarrhea of *Shigella* etiology in children presenting for diarrhea treatment. We showed how use of this clinical prediction rule in conjunction with emerging point of care diagnostic testing platforms could lead to major improvements in standardized, evidence-based care for diarrhea of *Shigella* etiology. *Shigella* remains a leading cause of diarrhea episodes and deaths for young children in LMICs, and improved tools for its management are critical for improving the health of children.

## Data Availability

All data is publicly available through clinepidb.org, and all code used for analysis is available from https://github.com/LeungLab/ShigellaDxStewardshipCPR.

https://clinepidb.org

## Notes

Funding information: This work was supported by National Institutes of Health under Ruth L. Kirschstein National Research Service Award NIH T32AI055434 and by the National Institute of Allergy and Infectious Diseases (R01AI135114).

The corresponding authors report no conflicts of interest for any authors.

### Competing Interest Statement

The authors have declared no competing interest.

### Funding Statement

This work was supported by National Institutes of Health under Ruth L. Kirschstein National Research Service Award NIH T32AI055434 and by the National Institute of Allergy and Infectious Diseases (R01AI135114).

### Author Declarations

Participants parents or caregivers provided informed consent, either in writing or witnessed if parents/caregivers were illiterate. The GEMS study protocol was approved by ethical review boards at each field site and the University of Maryland, Baltimore, USA. Informed consent was obtained from parents/caregivers of participants. The MAL-ED study protocol was approved by ethical review boards at each field site, the University of Virginia Institutional Review Board for Health Sciences Research, Charlottesville, USA, and the Johns Hopkins Institutional Review Board, Baltimore, USA.

## REFERENCES

1. Leibovici-Weissman Y, Neuberger A, Bitterman R, Sinclair D, Salam MA, Paul M. Antimicrobial drugs for treating cholera. Cochrane Database Syst Rev 2014; (6): CD008625.

2. Christopher PR, David KV, John SM, Sankarapandian V. Antibiotic therapy for Shigella dysentery. Cochrane Database Syst Rev 2010; (8): CD006784.

3. WHO. Integrated management of childhood illness: Chart Booklet. Geneva, Switzerland: World Health Organization, 2014.

4. WHO. Pocket book of Hospital care for children: guidelines for the management of common childhood illnesses. 2nd ed. Geneva, Switzerland: World Health Organization, 2013.

5. Pavia AT, Shipman LD, Wells JG, et al. Epidemiologic evidence that prior antimicrobial exposure decreases resistance to infection by antimicrobial-sensitive Salmonella. J Infect Dis 1990; 161(2): 255–60.

6. Gaufin T, Blumenthal J, Ramirez-Sanchez C, et al. Antimicrobial-Resistant Shigella spp. in San Diego, California, USA, 2017-2020. Emerg Infect Dis 2022; 28(6): 1110–6.

7. Sivapalasingam S, Nelson JM, Joyce K, Hoekstra M, Angulo FJ, Mintz ED. High prevalence of antimicrobial resistance among Shigella isolates in the United States tested by the National Antimicrobial Resistance Monitoring System from 1999 to 2002. Antimicrob Agents Chemother 2006; 50(1): 49–54.

8. Mamishi S, Pourakbari B, Ghaffari Charati M, Mahmoudi S, Abdolsalehi MR, Hosseinpour Sadeghi R. High frequency of antimicrobial resistance and virulence gene in Shigella species isolated from pediatric patients in an Iranian Referral Hospital. Acta Biomed 2022; 93(2): e2022027.

9. Pakbin B, Didban A, Bruck WM, Alizadeh M. Phylogenetic analysis and antibiotic resistance of Shigella sonnei isolates. FEMS Microbiol Lett 2022; 369(1).

10. Rostami A, Zadeh FA, Ebrahimzadeh F, Jafari-Sales A, Gholami S. Globally Vibrio cholera antibiotics resistance to RNA and DNA effective antibiotics: A systematic review and meta-analysis. Microb Pathog 2022: 105514.

11. Thuthikkadu Indhuprakash S, Karthikeyan M, Gopal G, et al. Antibody therapy against antibiotic-resistant diarrheagenic Escherichia coli: a systematic review. Immunotherapy 2021; 13(15): 1305–20.

12. Beyene AM, Gezachew M, Mengesha D, Yousef A, Gelaw B. Prevalence and drug resistance patterns of Gram-negative enteric bacterial pathogens from diarrheic patients in Ethiopia: A systematic review and meta-analysis. PLoS One 2022; 17(3): e0265271.

13. Kotloff KL, Nataro JP, Blackwelder WC, et al. Burden and aetiology of diarrhoeal disease in infants and young children in developing countries (the Global Enteric Multicenter Study, GEMS): a prospective, case-control study. Lancet 2013; 382(9888): 209–22.

14. Lindsay B, Ochieng JB, Ikumapayi UN, et al. Quantitative PCR for detection of Shigella improves ascertainment of Shigella burden in children with moderate-to-severe diarrhea in low-income countries. J Clin Microbiol 2013; 51(6): 1740–6.

15. Kotloff KL, Riddle MS, Platts-Mills JA, Pavlinac P, Zaidi AKM. Shigellosis. Lancet 2018; 391(10122): 801–12.

16. Tickell KD, Brander RL, Atlas HE, Pernica JM, Walson JL, Pavlinac PB. Identification and management of Shigella infection in children with diarrhoea: a systematic review and meta-analysis. Lancet Glob Health 2017; 5(12): e1235–e48.

17. Pavlinac PB, Denno DM, John-Stewart GC, et al. Failure of Syndrome-Based Diarrhea Management Guidelines to Detect Shigella Infections in Kenyan Children. J Pediatric Infect Dis Soc 2016; 5(4): 366–74.

18. Chakraborty S, Connor S, Velagic M. Development of a simple, rapid, and sensitive diagnostic assay for enterotoxigenic E. coli and Shigella spp applicable to endemic countries. PLoS Negl Trop Dis 2022; 16(1): e0010180.

19. Taneja N, Nato F, Dartevelle S, et al. Dipstick test for rapid diagnosis of Shigella dysenteriae 1 in bacterial cultures and its potential use on stool samples. PLoS One 2011; 6(10): e24830.

20. Connor S, Velagic M, Zhang X, et al. Evaluation of a simple, rapid and field-adapted diagnostic assay for enterotoxigenic E. coli and Shigella. PLoS Negl Trop Dis 2022; 16(2): e0010192.

21. Reilly BM, Evans AT. Translating clinical research into clinical practice: impact of using prediction rules to make decisions. Ann Intern Med 2006; 144(3): 201–9.

22. Kotloff KL, Blackwelder WC, Nasrin D, et al. The Global Enteric Multicenter Study (GEMS) of diarrheal disease in infants and young children in developing countries: epidemiologic and clinical methods of the case/control study. Clin Infect Dis 2012; 55 Suppl 4: S232–45.

23. Richard SA, Barrett LJ, Guerrant RL, Checkley W, Miller MA, Investigators M-EN. Disease surveillance methods used in the 8-site MAL-ED cohort study. Clin Infect Dis 2014; 59 Suppl 4: S220–4.

24. Platts-Mills JA, McCormick BJ, Kosek M, et al. Methods of analysis of enteropathogen infection in the MAL-ED Cohort Study. Clin Infect Dis 2014; 59 Suppl 4: S233–8.

25. Platts-Mills JA, Babji S, Bodhidatta L, et al. Pathogen-specific burdens of community diarrhoea in developing countries: a multisite birth cohort study (MAL-ED). Lancet Glob Health 2015; 3(9): e564–75.

26. Investigators M-EN. The MAL-ED study: a multinational and multidisciplinary approach to understand the relationship between enteric pathogens, malnutrition, gut physiology, physical growth, cognitive development, and immune responses in infants and children up to 2 years of age in resource-poor environments. Clin Infect Dis 2014; 59 Suppl 4: S193–206.

27. Liu J, Platts-Mills JA, Juma J, et al. Use of quantitative molecular diagnostic methods to identify causes of diarrhoea in children: a reanalysis of the GEMS case-control study. Lancet 2016; 388(10051): 1291–301.

28. James G, Witten D, Hastie T, Tibshirani R. An introduction to statistical learning with applications in R. New York: Springer, 2013.

29. Van Calster B, McLernon DJ, van Smeden M, et al. Calibration: the Achilles heel of predictive analytics. BMC Med 2019; 17(1): 230.

30. Steyerberg EW, Vergouwe Y. Towards better clinical prediction models: seven steps for development and an ABCD for validation. Eur Heart J 2014; 35(29): 1925–31.

31. Tickell KD, Mangale DI, Tornberg-Belanger SN, et al. A mixed method multi-country assessment of barriers to implementing pediatric inpatient care guidelines. PLoS One 2019; 14(3): e0212395.

32. Lassi ZS, Middleton PF, Bhutta ZA, Crowther C. Strategies for improving health care seeking for maternal and newborn illnesses in low- and middle-income countries: a systematic review and meta-analysis. Glob Health Action 2016; 9: 31408.

