## Supplemental File for "Clinical prediction rule to guide diagnostic testing for *Shigellosis* and improve antibiotic stewardship for pediatric diarrhea"

Additional methods for assessing model calibration: We assessed calibration intercept, or calibration-in-the-large, by modeling the log-odds of the true status, offset by the CPR-predicted log-odds. In other words, we used logistic regression to estimate the mean while subtracting out the estimate. Next, we fit a logistic regression model with the CPR-predicted log-odds as the independent variable and the log-odds of the true status as the dependent variable, in order to calculate the calibration slope and assess the spread of the estimated probabilities. Finally, we graphically assessed moderate calibration. We calculated the predicted probability of *Shigella* infection for each child in a given analysis using each iteration of each n-variable model fit. We then binned these predicted probabilities into deciles, and calculated the proportion of each decile who truly experienced the outcome for each iteration of each n-variable model. We then calculated the mean predicted probability and observed proportion for each decile across iterations, averaged these observed proportions, and plotted them against averaged deciles for each n-variable model fit.

Figure S1: Schematic of CPR-guided diagnostic testing regimen

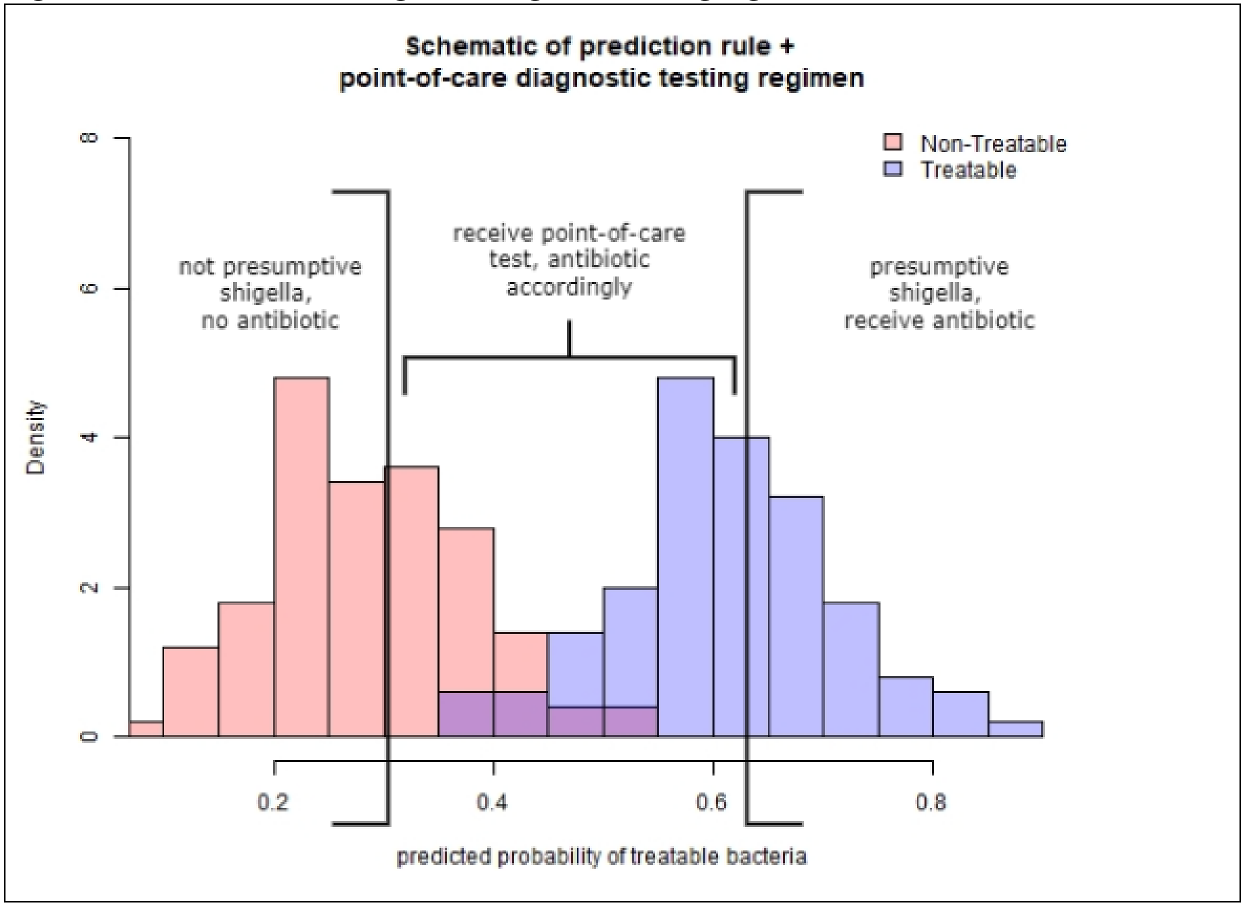

Figure S2: Flow diagram of study inclusion

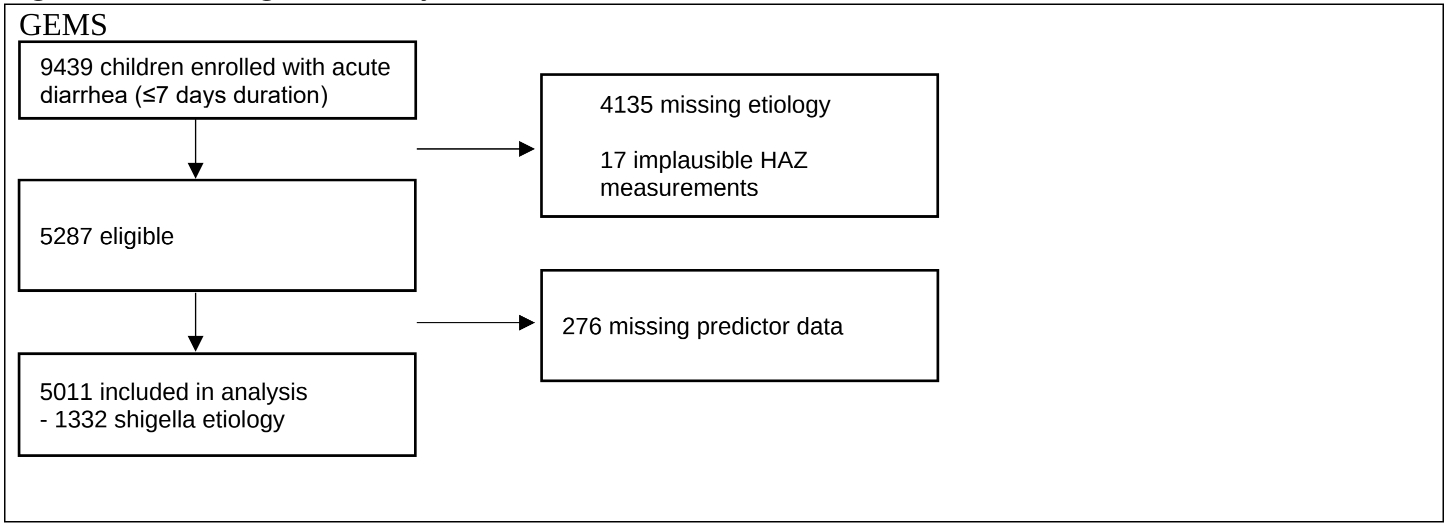

Table S1: Full list of considered predictor variables

| |
|---|
| Study site (site) |
| Child sex (f3_gender) |
| Loss of skin turgor (f3_drh_turgor) |
| Intravenous rehydration (f3_drh_iv) |
| Hospitalized (f3_drh_hosp) |
| Your relationship to the child (f4a_relationship) |
| Where child's father lives (f4a_dad_live) |
| Primary caregiver's max school (f4a_prim_schl) |
| People living in house 6 months (f4a_ppl_house) |
| Children under 60 months in the house (f4a_yng_children) |
| How many rooms used for sleeping (f4a_slp_rooms) |
| Predominant floor (f4a_floor) |
| Electricity (f4a_house_elec) |
| Bicycle/rickshaw (f4a_house_bike) |
| Telephone (f4a_house_phone) |
| Television (f4a_house_tele) |
| Car/truck (f4a_house_car) |
| Animal-drawn cart (f4a_house_cart) |
| Motorcycle/scooter (f4a_house_scoot) |
| Refrigerator (f4a_house_fridge) |
| Agricultural land (f4a_house_agland) |
| Radio (f4a_house_radio) |
| Boat with motor (f4a_house_boat) |
| None of the above assets (f4a_house_none) |
| Electricity (f4a_fuel_elec) |
| Biogas (f4a_fuel_biogas) |
| Straw/shrubs/grass (f4a_fuel_grass) |
| Liquid propane gas (f4a_fuel_propane) |
| Coal/lignite (f4a_fuel_coal) |
| Animal dung (f4a_fuel_dung) |
| Natural gas (f4a_fuel_natgas) |
| Charcoal (f4a_fuel_charcoal) |

Agricultural crop residue (f4a_fuel_crop)
Kerosene (f4a_fuel_kero)
Wood (f4a_fuel_wood)
Other fuel (f4a_fuel_other)
Goat (f4a_ani_goat)
Sheep (f4a_ani_sheep)
Dog (f4a_ani_dog)
Cat (f4a_ani_cat)
Cow (f4a_ani_cow)
Rodents (f4a_ani_rodents)
Fowl (f4a_ani_fowl)
Other animal (f4a_ani_other)
No animals (f4a_ani_no)
Water piped to house (f4a_water_house)
Covered well in house/yard (f4a_water_covwell)
Water piped into yard (f4a_water_yard)
Covered public well (f4a_water_covpwell)
Public tap (f4a_water_pubtap)
Protected spring (f4a_water_prospring)
Open well in house/yard (f4a_water_well)
Unprotected spring (f4a_water_unspring)
Open public well (f4a_water_pubwell)
River/stream (f4a_water_river)
Pond/lake (f4a_water_pond)
Deep tube well (f4a_water_deepwell)
Rainwater (f4a_water_rain)
Shallow tube well (f4a_water_shallwell)
Bought water (f4a_water_bought)
Other water source (f4a_water_othr)
Bore hole (f4a_water_bore)
Main source of drinking water (f4a_ms_water*)
How often is water available (f4a_water_avail)
Did you give the child stored water (f4a_store_water)

| |
|---|
| Do you usually treat drinking water? (f4a_trt_water)<br>Usual treatment method (f4a_trt_method)<br>How are child’s feces disposed (f4a_disp_feces)<br>Facility used to dispose of feces (f4a_fac_waste)<br>How many households share facility? (f4a_share_fac)<br>Wash hands before eating? (f4a_wash_eat)<br>Wash hands before cooking (f4a_wash_cook)<br>Wash hands before you nurse? (f4a_wash_nurse)<br>Wash hands after you defecate (f4a_wash_def)<br>Wash hands after handling animals (f4a_wash_animal)<br>Wash hands after cleaning a child (f4a_wash_child)<br>Wash hands other times (f4a_wash_othr)<br>What do you use to wash your hands? (f4a_wash_use)<br>Is the child currently breastfed? (f4a_breastfed)<br>How long as this diarrhea episode lasted (days)? (f4a_drh_days)<br>Maximum number of loose stools (f4a_max_stools)<br>Caregiver reported blood in stools (f4a_drh_blood)<br>Vomiting 3 or more times per day (f4a_drh_vomit)<br>Very thirsty (f4a_drh_thirst)<br>Drank much less than usual (f4a_drh_lessdrink)<br>Belly pain (f4a_drh_bellypain)<br>Irritable or restless (f4a_drh_restless)<br>Decreased activity or lethargy (f4a_drh_lethrgy)<br>Loss of consciousness (f4a_drh_consc)<br>Rectal straining (f4a_drh_strain)<br>Rectal prolapse (f4a_drh_prolapse)<br>Cough (f4a_drh_cough)<br>Convulsions (f4a_drh_conv)<br>Very thirsty (f4a_cur_thirsty)<br>Wrinkled skin (f4a_cur_skin)<br>Irritable or restless (f4a_cur_restless)<br>Dry mouth (f4a_cur_drymouth)<br>Fast breathing (f4a_cur_fastbreath) |

| |
|---|
| ORALITE or ORS (f4a_hometrt_ors)<br>Homemade fluid (f4a_hometrt_maize)<br>Special mile or infant formula (f4a_hometrt_milk)<br>Home remedy/herbal medication (f4a_hometrt_herb)<br>Zinc (f4a_hometrt_zinc)<br>No special remedies given (f4a_hometrt_none)<br>Any other liquids (f4a_hometrt_othrliq)<br>Antibiotics (f4a_hometrt_ab)<br>Other treatment (f4a_hometrt_othr1)<br>Other treatment (f4a_hometrt_othr2)<br>How much offered to drink (f4a_offr_drink)<br>Seek outside care (f4a_seek_outside)<br>Pharmacy (f4a_seek_pharm)<br>Friend/relative (f4a_seek_friend)<br>Traditional healer (f4a_seek_healer)<br>Unlicensed practitioner (f4a_seek_doc)<br>Licensed practitioner (f4a_seek_privdoc)<br>Bought a remedy (f4a_seek_remdy)<br>Other hospital/center (f4a_seek_other)<br>Mid-upper arm circumference (f4b_muac)<br>Axillary temperature (f4b_temp)<br>Respiratory rate per minute (f4b_resp)<br>Chest indrawing (f4b_chest_indrw)<br>Eyes (f4b_eyes)<br>Mouth (f4b_mouth)<br>Skin pinch (f4b_skin)<br>Mental status (f4b_mental)<br>Rectal prolapse (f4b_rectal)<br>Bipedal edema (f4b_bipedal)<br>Abnormal hair (f4b_abn_hair)<br>Undernutrition (f4b_under_nutr)<br>Skin as ‘flaky paint’ appearance (f4b_skin_flaky)<br>Receive rehydration here (f4b_recommend) |

| |
|---|
| Child was admitted to hospital (f4b_admit) |
| Child age (months) (base_age) |

*f4a_ms_water recategorized into the following: surface, other unimproved, other improved, piped, other(1, 2)

Table S2: Total sample size and etiology by site

| GEMS<br><br>N (%) | The Gambia<br>682 | Mali<br>833 | Mozambique<br>479 | Kenya<br>786 | India<br>849 | Bangladesh<br>876 | Pakistan<br>782 |
|---|---|---|---|---|---|---|---|
| Shigella | | | | | | | |
| AFe ≥ 0.3 | 194 (28.45) | 184 (22.09) | 107 (22.34) | 142 (18.07) | 194 (22.85) | 500 (57.08) | 261 (33.38) |
| AFe ≥ 0.5 | 160 (23.46) | 149 (17.89) | 96 (20.04) | 104 (13.23) | 162 (19.08) | 496 (56.62) | 223 (28.52) |
| AFe ≥ 0.7 | 121 (17.74) | 99 (11.88) | 86 (17.95) | 66 (8.40) | 118 (13.90) | 486 (55.48) | 171 (21.87) |

Table S3: Total sample size and etiology by age

| GEMS<br><br>N (%) | Total<br><br>5287 | 0-11mo<br><br>1904 | 12-23mo<br><br>1828 | 24-59mo<br><br>1555 |
|---|---|---|---|---|
| Shigella | | | | |
| AFe ≥ 0.3 | 1582 (29.92%) | 201 (10.56) | 718 (39.28) | 663 (42.64) |
| AFe ≥ 0.5 | 1390 (26.29%) | 165 (8.67) | 643 (35.18) | 582 (37.43) |
| AFe ≥ 0.7 | 1147 (21.69%) | 115 (6.04) | 526 (28.77) | 506 (32.54) |
| MAL-ED | 1226 | 679 | 547 | |
| Shigella | | | | |
| AFe ≥ 0.5 | 153 (12.48%) | 26 (3.8%) | 127 (23.2%) | |

Figure S3: Number of variables and AUC for random forest regression and logistic regression

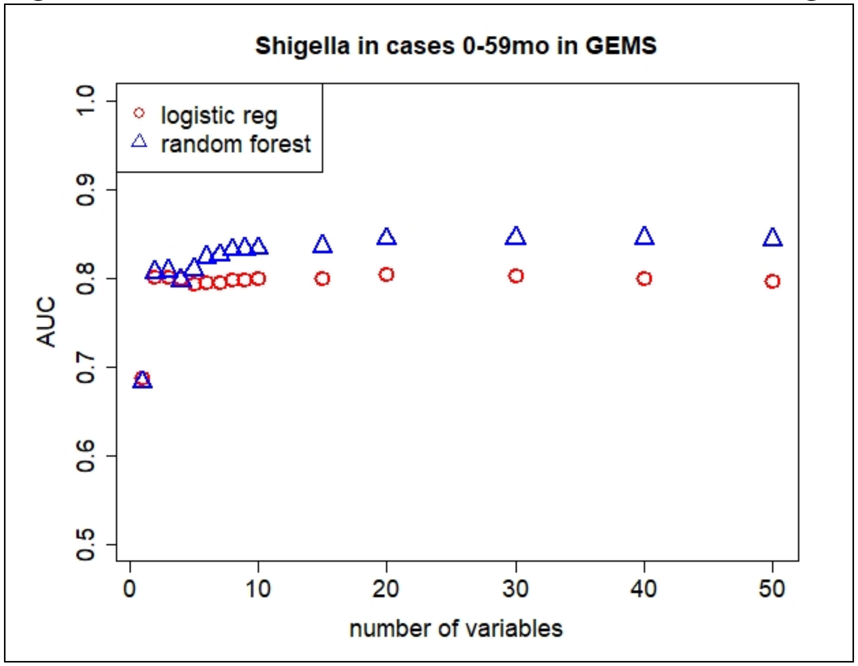

Figure S4: Distributions of top predictive variables in GEMS and MAL-ED datasets

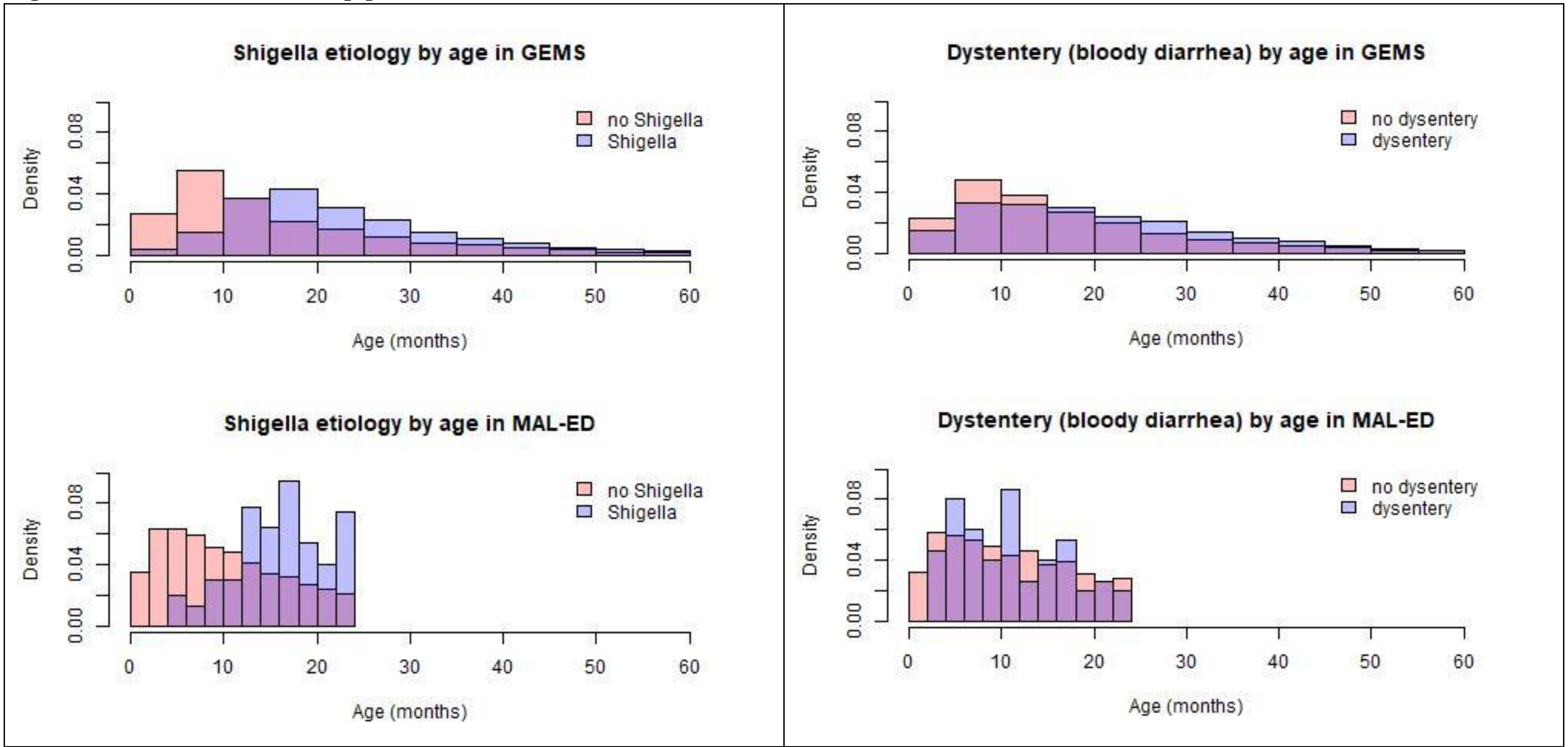

| GEMS | Bloody diarrhea |
|---|---|
| Shigella AFe ≥ 0.5 n=1332 | 775 (58.2%) |

Table S4: Cross-validated average AUC, odds ratios, and 95% confidence intervals for logistic regression model predicting shigella etiology (AFe $\geq 0.5$) in children 0-59mo in LMICs in GEMS

| GEMS | |
|---|---|
| AUC (95% CI): | 0.80 (0.79, 0.82) |
| Variables | OR (95% CI) |
| Age (months) | 1.04 (1.03, 1.04) |
| Caregiver reported blood in stool | 9.53 (8.21, 11.09) |

Table S5: Variable importance ordering and cross-validated average overall AUC and AUC by patient subset and 95% confidence intervals for a 2 (bold), 5 (plain text), and 10 (italicized) variable logistic regression model for predicting shigella etiology. Models were derived using all children of all ages 0-59mo from all sites in GEMS, and defined shigella etiology as AFe ≥ 0.5, unless otherwise noted.

| Patient subset | Only MUAC considered (main text model) | Only HAZ considered (not MUAC) | HAZ + MUAC considered | 0-11mo | 12-23mo | 0-23mo | 24-59mo |
|---|---|---|---|---|---|---|---|
| **AUCs – 2 variable** | **0.80 (0.80, 0.81)** | **0.80 (0.80, 0.81)** | **0.80 (0.80, 0.81)** | **0.74 (0.73, 0.75)** | **0.72 (0.72, 0.73)** | **0.82 (0.82, 0.82)** | **0.80 (0.79, 0.81)** |
| 5 | 0.79 (0.79, 0.80) | 0.79 (0.79, 0.80) | 0.80 (0.80, 0.81) | 0.78 (0.77, 0.79) | 0.73 (0.73, 0.74) | 0.82 (0.82, 0.83) | 0.82 (0.82, 0.83) |
| 10 | *0.80 (0.80, 0.80)* | *0.80 (0.79, 0.80)* | *0.80 (0.80, 0.80)* | *0.77 (0.76, 0.78)* | *0.76 (0.76, 0.77)* | *0.83 (0.82, 0.83)* | *0.82 (0.81, 0.82)* |
| 1 | Age (months) | Age (months) | Age (months) | Age (months) | Caregiver reported blood in stool | Age (months) | Caregiver reported blood in stool |
| 2 | Caregiver reported blood in stool | Caregiver reported blood in stool | Caregiver reported blood in stool | Caregiver reported blood in stool | MUAC | Caregiver reported blood in stool | Eyes |
| 3 | MUAC | HAZ | HAZ | MUAC | Respiratory rate | Respiratory rate | Site |
| 4 | Respiratory rate | Respiratory rate | MUAC | Respiratory rate | Temperature | MUAC | MUAC |
| 5 | Temperature | Temperature | Respiratory rate | Temperature | Age (months) | Temperature | Temperature |
| 6 | Sunken eyes | Sunken eyes | Temperature | Num. people living in household | Sunken eyes | Num. people living in household | Respiratory rate |

| | | | | | | | |
|---|---|---|---|---|---|---|---|
| 7 | Num. people living in household | Num. people living in household | Sunken eyes | Num. days of diarrhea at presentation | Num. people living in household | Sunken eyes | Straw/shrubs /grass used for fuel |
| 8 | Site | Site | Num. people living in household | Num. rooms used for sleeping | Site | Breastfed | Age |
| 9 | Num. days of diarrhea at presentation | Num. days of diarrhea at presentation | Site | How many households share facility | Num. days of diarrhea at presentation | Num. days of diarrhea at presentation | Num. people living in household |
| 10 | Num. rooms used for sleeping | Num. rooms used for sleeping | Num. days of diarrhea at presentation | How much offered to drink since diarrhea | Rectal straining | Num. rooms used for sleeping | Did you give the child stored water |

| Patient subset | AFe ≥ 0.3 | AFe ≥ 0.7 | Only fit to cases reporting bloody diarrhea (dysentery) | Only fit to cases not reporting bloody diarrhea (no dysentery) | All sites except Bangladesh | Also consider clinically observed stool descriptors | Also consider season of diarrhea (Apr-Sept) |
|---|---|---|---|---|---|---|---|
| **AUCs – 2 variable** | **0.79 (0.79, 0.79)** | **0.82 (0.82, 0.83)** | **0.73 (0.72, 0.74)** | **0.66 (0.65, 0.66)** | **0.75 (0.74, 0.75)** | **0.80 (0.80, 0.81)** | **0.80 (0.80, 0.81)** |
| 5 | 0.78 (0.78, 0.78) | 0.82 (0.82, 0.83) | 0.73 (0.73, 0.74) | 0.64 (0.64, 0.65) | 0.74 (0.73, 0.74) | 0.80 (0.80, 0.81) | 0.79 (0.79, 0.80) |
| 10 | *0.78 (0.78, 0.78)* | *0.82 (0.82, 0.83)* | *0.74 (0.74, 0.75)* | *0.64 (0.63, 0.64)* | *0.73 (0.73, 0.74)* | *0.80 (0.80, 0.81)* | *0.80 (0.80, 0.80)* |
| 1 | Age (months) | Age (months) | Age (months) | Age (months) | Caregiver reported blood in stool | Age (months) | Age (months) |

| | | | | | | | |
|---|---|---|---|---|---|---|---|
| 2 | Caregiver reported blood in stool | Caregiver reported blood in stool | Temperature | MUAC | Age (months) | Caregiver reported blood in stool | Caregiver reported blood in stool |
| 3 | MUAC | Eyes | Respiratory rate | Respiratory rate | MUAC | Clinician observed blood in stool | MUAC |
| 4 | Respiratory rate | MUAC | MUAC | Temperature | Respiratory rate | MUAC | Respiratory rate |
| 5 | Temperature | Respiratory rate | Num. people living in household | Num. people living in household | Temperature | Respiratory rate | Temperature |
| 6 | Sunken eyes | Temperature | Num. days of diarrhea at presentation | Num. days of diarrhea at presentation | Num. people living in household | Temperature | Sunken eyes |
| 7 | Num. people living in household | Site | Num. rooms used for sleeping | Num. rooms used for sleeping | Num. days of diarrhea at presentation | Sunken eyes | Num. people living in household |
| 8 | Num. days of diarrhea at presentation | Num. people living in household | Maximum number of loose stools | How many households share facility | Num. rooms used for sleeping | Num. people living in household | Site |
| 9 | Site | Rectal straining | How many households share facility? | Num. children <60 months live in household | How many households share facility? | Site | Num. days of diarrhea at presentation |
| 10 | Num. rooms used for sleeping | Straw/shrubs /grass used for fuel | Primary caregiver's max school | breastfed | Num. children <60 months live in household | Consistency | Num. rooms used for sleeping |
| Ranking of addition | | | | | | 18 – mucus<br>84 – pus | 30 – season |

| |
|---|
| ally considered variables |

| Patient subset | The Gambia | Mali | Mozambique | Kenya | India | Bangladesh | Pakistan | Fit on data from African countries | Fit on data from Asian countries, excluding Bangladesh |
|---|---|---|---|---|---|---|---|---|---|
| **AUCs – 2 variable** | **0.78 (0.77, 0.79)** | **0.63 (0.62, 0.64)** | **0.82 (0.81, 0.83)** | **0.61 (0.60, 0.63)** | **0.80 (0.80, 0.81)** | **0.85 (0.84, 0.86)** | **0.74 (0.73, 0.75)** | **0.74 (0.74, 0.75)** | **0.76 (0.76, 0.77)** |
| 5 | 0.77 (0.76, 0.78) | 0.60 (0.59, 0.61) | 0.82 (0.81, 0.83) | 0.68 (0.66, 0.69) | 0.79 (0.78, 0.80) | 0.90 (0.89, 0.90) | 0.71 (0.70, 0.72) | 0.72 (0.71, 0.73) | 0.75 (0.74, 0.76) |
| 10 | *0.78 (0.77, 0.79)* | *0.57 (0.56, 0.58)* | *0.81 (0.80, 0.83)* | *0.67 (0.65, 0.68)* | *0.77 (0.76, 0.78)* | *0.91 (0.90, 0.91)* | *0.72 (0.71, 0.73)* | *0.73 (0.72, 0.73)* | *0.75 (0.74, 0.75)* |
| 1 | Caregiver reported blood in stool | Age (months) | Age (months) | Age (months) | Caregiver reported blood in stool | Age (months) | Caregiver reported blood in stool | Age (months) | Caregiver reported blood in stool |
| 2 | Age (months) | MUAC | Caregiver reported blood in stool | Respiratory rate | Age (months) | Temperature | Age (months) | Caregiver reported blood in stool | Age (months) |
| 3 | MUAC | Respiratory rate | MUAC | MUAC | MUAC | MUAC | MUAC | Respiratory rate | MUAC |
| 4 | Num. people living in household | Temperature | Respiratory rate | Caregiver reported blood in stool | Respiratory rate | Caregiver reported blood in stool | Respiratory rate | MUAC | Respiratory rate |
| 5 | Respiratory rate | Num. people living in household | Breastfed | Temperature | Temperature | Respiratory rate | Temperature | Temperature | Temperature |

| | | | | | | | | | |
|---|---|---|---|---|---|---|---|---|---|
| 6 | Temperature | Num. rooms used for sleeping | Temperature | Num. people living in household | How many households share facility | Sunken eyes | Num. people living in household | Num. people living in household | Num. people living in household |
| 7 | Num. rooms used for sleeping | Num. children <60 months live in household | Num. people living in household | How many households share facility | Num. people living in household | Rectal straining | Num. days of diarrhea at presentation | Num. rooms used for sleeping | How many households share facility? |
| 8 | Num. children <60 months live in household | Num. days of diarrhea at presentation | Vomiting 3 or more times per day | Num. days of diarrhea at presentation | Num. days of diarrhea at presentation | Num. days of diarrhea at presentation | Num. children <60 months live in household | Num. days of diarrhea at presentation | Num. days of diarrhea at presentation |
| 9 | Sunken eyes | How many households share facility | admit | Primary caregiver's max school | Primary caregiver's max school | Num. people living in household | Received rehydration here | Num. children <60 months live in household | Num. children <60 months live in household |
| 10 | Received rehydration here | Primary caregiver's max school | Num. days of diarrhea at presentation | Maximum number of stools per day | Child feces disposal | Belly pain | How often water available at main source in past 2 weeks | Breastfed | Primary caregiver's max school |
| | | | | | | | | 2-variable CPR performance in data from Asia (excluding Bangladesh) | 2-variable CPR performance in data from Africa |

| | | | | | | | | | |
|---|---|---|---|---|---|---|---|---|---|
| | | | | | | | | 0.77 (0.74, 0.80) | 0.74 (0.71, 0.76) |

Unless otherwise noted, only MUAC was considered as a potential predictor, not HAZ.

Figure S5: Calibration Curves

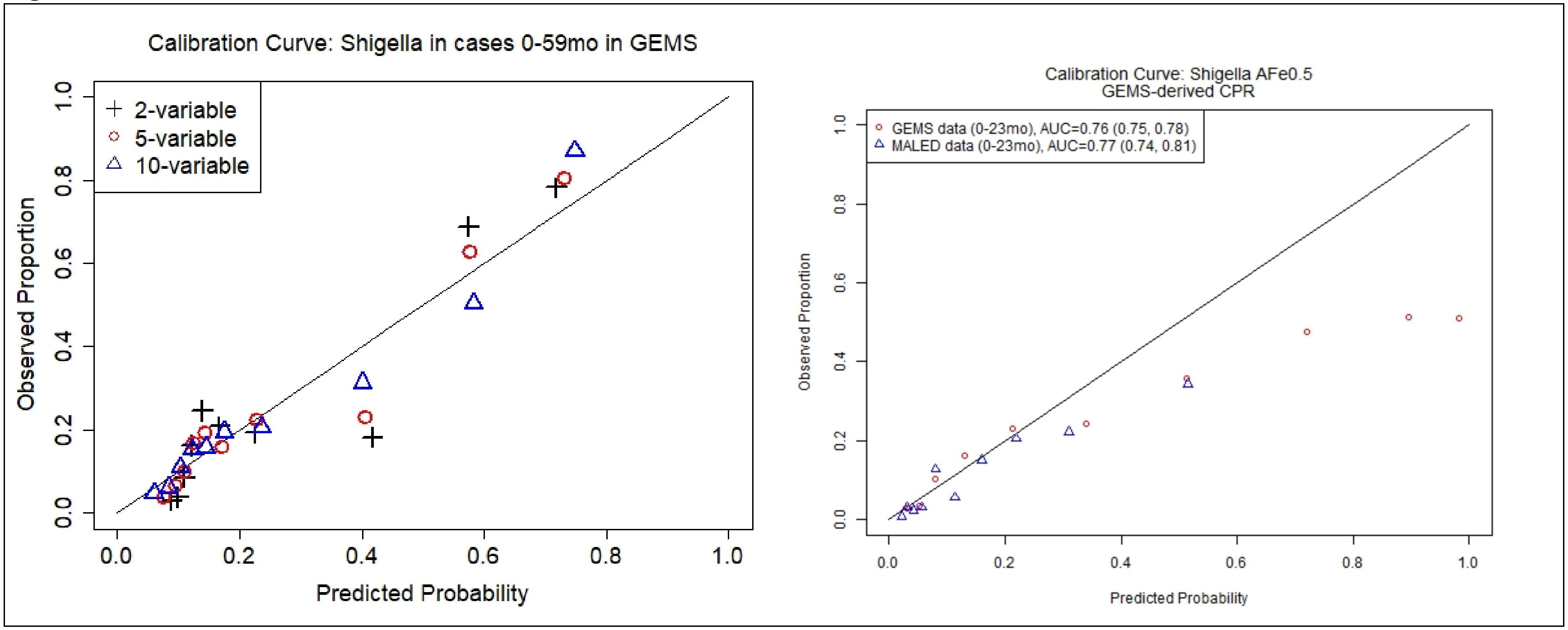

Figure S6: Impact of POC test accuracy on performance of CPR-guided diagnostic testing regimen.

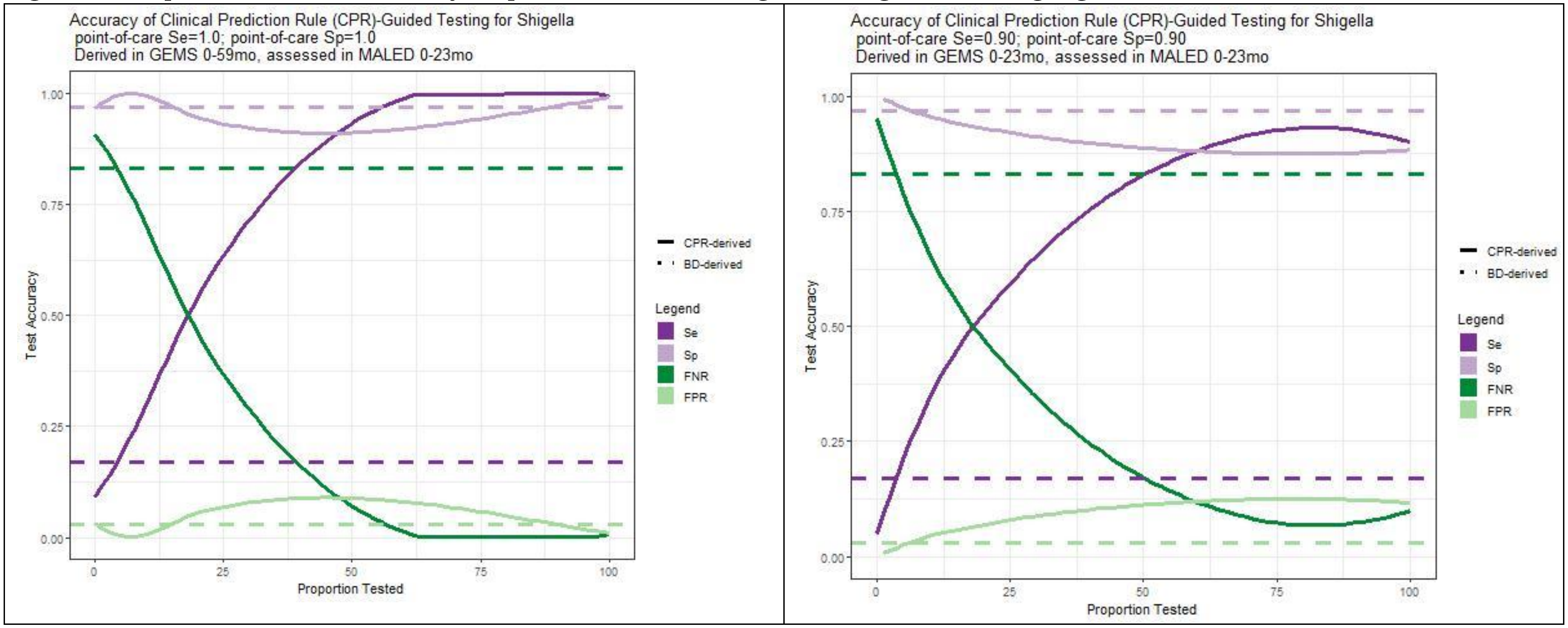

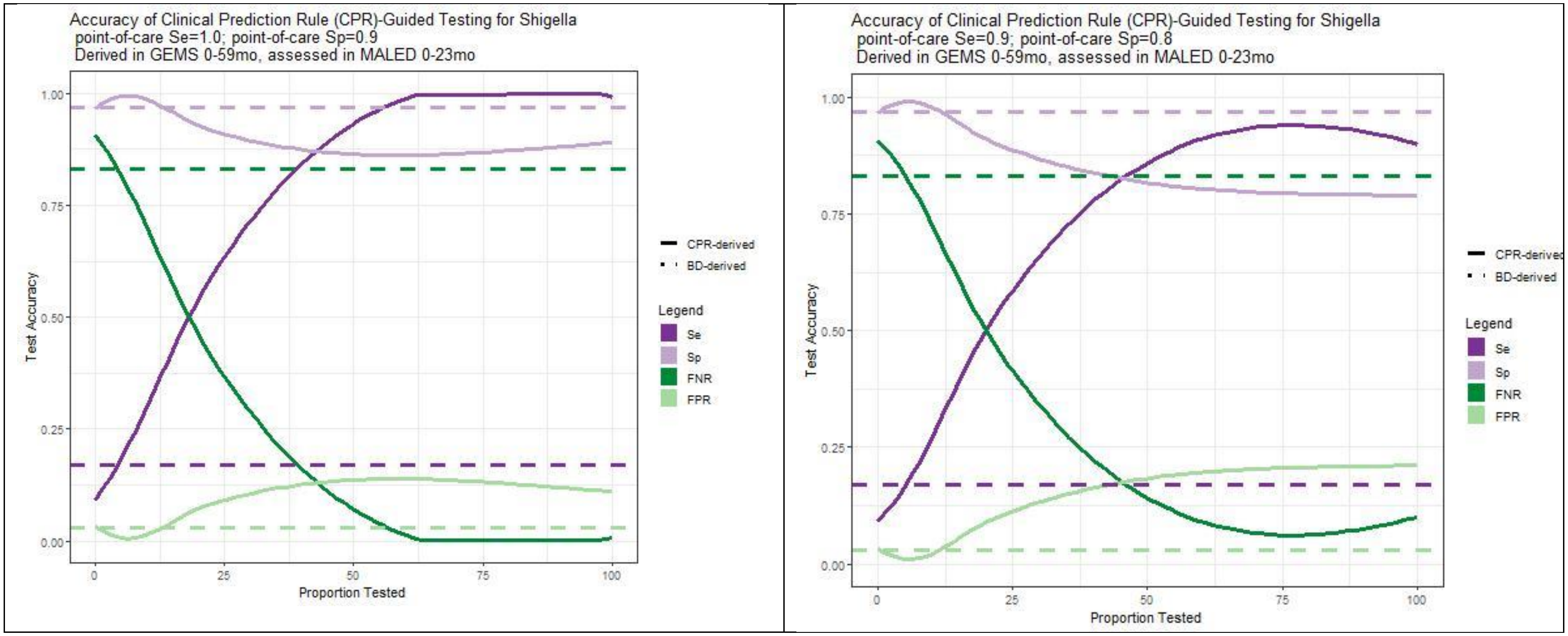

Accuracy of Clinical Prediction Rule (CPR)-Guided Testing for Shigella
point-of-care Se=1.0; point-of-care Sp=0.9
Derived in GEMS 0-59mo, assessed in MALED 0-23mo
Test Accuracy
Proportion Tested
CPR-derived
BD-derived
Legend
Se
Sp
FNR
FPR
Accuracy of Clinical Prediction Rule (CPR)-Guided Testing for Shigella
point-of-care Se=0.9; point-of-care Sp=0.8
Derived in GEMS 0-59mo, assessed in MALED 0-23mo
Test Accuracy
Proportion Tested
CPR-derived
BD-derived
Legend
Se
Sp
FNR
FPR

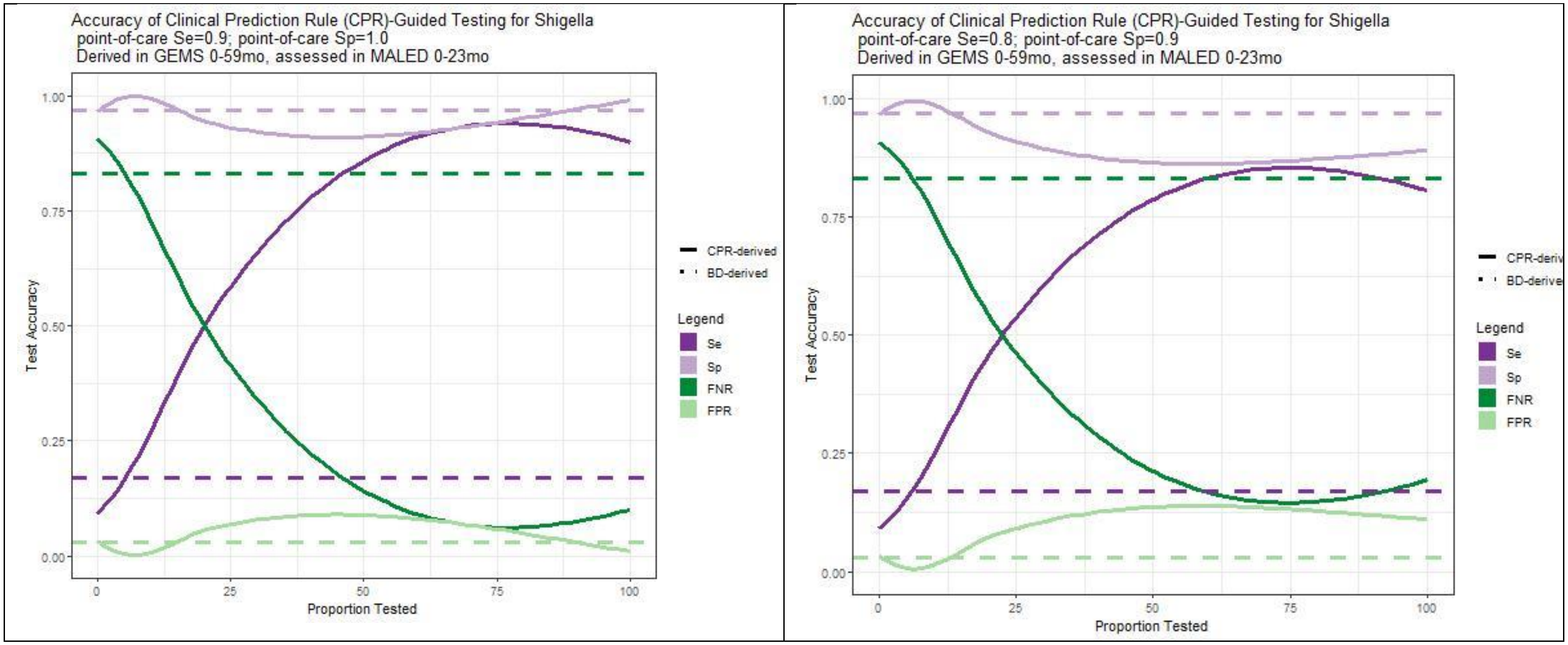
Accuracy of Clinical Prediction Rule (CPR)-Guided Testing for Shigella
point-of-care Se=0.9; point-of-care Sp=1.0
Derived in GEMS 0-59mo, assessed in MALED 0-23mo
Test Accuracy
1.00
0.75
0.50
0.25
0.00
0
25
50
75
100
Proportion Tested
CPR-derived
BD-derived
Legend
Se
Sp
FNR
FPR
Accuracy of Clinical Prediction Rule (CPR)-Guided Testing for Shigella
point-of-care Se=0.8; point-of-care Sp=0.9
Derived in GEMS 0-59mo, assessed in MALED 0-23mo
Test Accuracy
1.00
0.75
0.50
0.25
0.00
0
25
50
75
100
Proportion Tested
CPR-deriv
BD-derive
Legend
Se
Sp
FNR
FPR

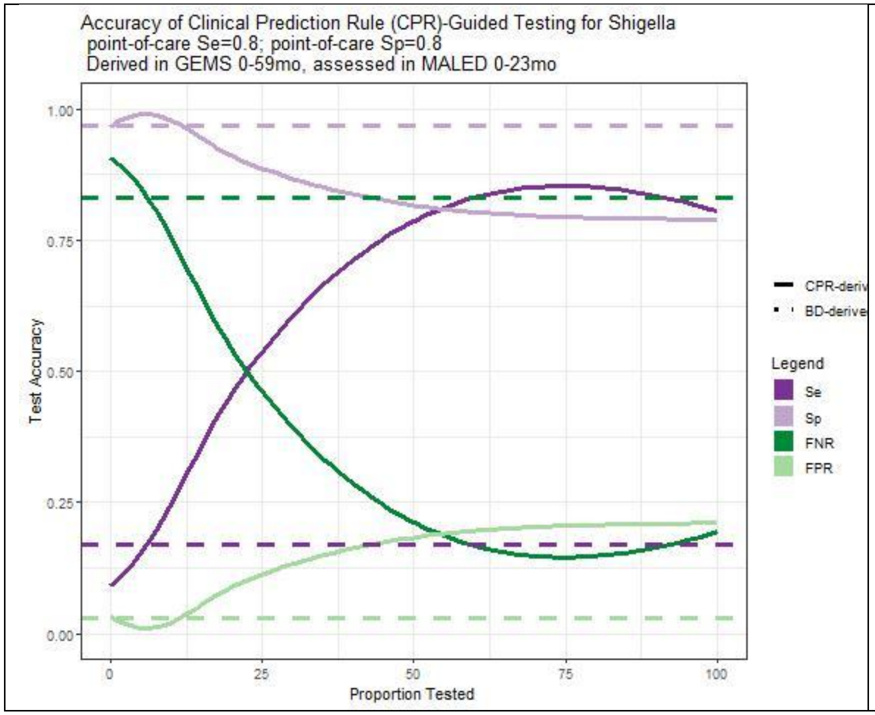
Accuracy of Clinical Prediction Rule (CPR)-Guided Testing for Shigella
point-of-care Se=0.8; point-of-care Sp=0.8
Derived in GEMS 0-59mo, assessed in MALED 0-23mo
1.00
0.75
0.50
0.25
0.00
Test Accuracy
0
25
50
75
100
Proportion Tested
CPR-deriv
BD-derive
Legend
Se
Sp
FNR
FPR
